# Re-evaluation of whole exome sequencing, including intronic region, in combination with genetic intolerance score for detecting foetal structural anomalies in X-linked disorders

**DOI:** 10.1101/2023.02.05.23285039

**Authors:** Kosuke Taniguchi, Fuyuki Hasegawa, Yuka Okazaki, Asuka Hori, Hiroko Ogata-Kawata, Saki Aoto, Osuke Migita, Tomoko Kawai, Kazuhiko Nakabayashi, Kohji Okamura, Kana Fukui, Seiji Wada, Katsusuke Ozawa, Yushi Ito, Haruhiko Sago, Kenichiro Hata

## Abstract

**Background:** Whole-exome sequencing (WES) is a strong diagnostic tool for foetal structural anomalies, but the causative gene for more than half the anomalies have not been identified. Therefore, improving the diagnostic yield based on WES data is essential.

**Methods:** First, 138 foetuses with structural anomalies were assessed using conventional WES and copy number variation (CNV) analyses. For undiagnosed cases, we employed a three-step approach for diagnosis. We re-evaluated 1) candidate variants using a loss-of-function observed/expected upper bound fraction (LOEUF) score, 2) all variants of disease-causing genes for clinically diagnosed cases using spliceAI, and 3) the rare variants in all low LOEUF scored genes (< 0.35) using spliceAI.

**Results:** We identified molecular diagnoses in 53 of 138 cases (38.4%) using conventional WES and CNV. For undiagnosed cases, for the first step, we diagnosed two X-linked recessive diseases. For the second step, we diagnosed Meckel-Gruber syndrome by detecting likely pathogenic intron variant in *TMEM67*. In the third step, we identified a *de novo* hemizygous pathogenic variant in one severe hydrops fetalis male, which caused aberrant splicing in *CASK*. We found a novel phenotype, hydrops fetalis, in CASK-related X-linked dominant disorder. Moreover, we revealed that the LOEUF score of X-linked disease-causing genes was significantly lower than that of autosomal genes among all OMIM-registered genes.

**Conclusion:** We showed that the evaluation of variants, including introns of WES data, in combination with the LOEUF score, could improve the WES diagnostic yield and be useful for evaluation of variants, especially on chromosome X.

**What is already known on this topic?:** Molecular genetic diagnosis of foetal structural anomalies using WES is being increasingly implemented. However, more than half of the cases cannot be diagnosed. There is signigicant potential to increase the diagnostic yield by re-analysing WES data.

**What this study adds:** In the present study, we focused on loss-of-function observed/expected upper bound fraction (LOEUF) scores to quantify genetic intolerance, and additional intron analysis for undiagnosed cases using conventional WES data. These approaches enabled the appropriate evaluation of candidate variants and detected overlooked candidate variants on intron. We diagnosed two X-linked recessive disorder cases (Hardikar syndrome and Ritscher-Schinzel syndrome), by re-evaluating candidate variants using the LOEUF score. We also diagnosed one Meckel-Gruber syndrome case caused by an intronic pathogenic variant, that had been overlooked by the conventional method. Moreover, evaluating all variants, including introns with low LOEUF score genes (2,971 genes) that could cause haploinsufficiency helped us find a pathogenic intronic variant on *CASK* in one hydrops fetalis case, which revealed that CASK-related X-linked dominant disorder could cause hydrops fetalis as severe phenotypes. Finally, the LOEUF score of X-linked genes was significantly lower than that of autosomal genes among all OMIM-registered genes, which meant that gene evaluation using the LOEUF score was helpful, for genetic diagnosis, especially for genes on chromosome X.

**How this study might affect research, practice, or policy:** The evaluation of variants, including introns, in combination with the LOEUF score is expected to contribute to the improvement of the diagnostic yield in WES. These approaches are easy and convenient to implement. The LOEUF score might be useful for evaluation of variants, especially in chromosome X.

## INTRODUCTION

The widespread use of next-generation sequencing (NGS) has revolutionised the available options for prenatal diagnosis.[1 2] Ultrasonography is a highly sensitive, cost-effective, and widely used diagnostic tool for congenital anomalies.[3] Accurate diagnosis of congenital anomalies is critical for the accurate assessment of prognosis and development of potential treatment during pregnancy or after birth and risk of recurrence in future pregnancies.[1] Recent advances in NGS have enabled the precise determination and validation of the accuracy of ultrasonography, particularly, to detect congenital genetic risks and defects. As genomic analysis for prenatal diagnosis may become more widespread, it is essential to determine whether the retrieved variants are pathogenic. Genomic analysis of foetal-onset structural anomalies could provide a broader range of variant information and enrich disease genomic databases.

Recently, there has been an increase in comprehensive analyses employing whole-exome sequencing (WES) for foetal structural anomalies and stillbirths.[1 2 4 5] The genetic diagnostic yield for foetal structural anomalies is approximately 10%–20%.[1 2 5] Although there are many cases where the cause cannot be identified, clinical applications of perinatal comprehensive genetic testing are considered.[6 7]

Genes with fewer functional variants in healthy individuals are more likely to cause disorders than those with more functional variants.[8] A gene is ‘intolerant’ if there are relatively fewer genetic variants than expected in the general population.[9] Accordingly, here, we used the loss-of-function (LOF) observed/expected upper bound fraction (LOEUF) score for each gene, which was upgraded compared to the pLI (LOF intolerant) scores. Genes with low LOEUF scores could cause haploinsufficiency. In 2020, a WES study of a stillbirth cohort reported that stillbirth cases are enriched in LOF variants (nonsense, splicing abnormalities, and frameshift) in genes with low LOEUF scores, and some of which may be unknown, disease-causing genes.[5] Therefore, evaluation using the LOEUF scores might help us have new inslight to foetal onset diseases.

Recent studies reported that WES re-analysis with the latest pipeline might improve diagnostic yield for undiagnosed Mendelian diseases.[10-12] A previous study reported that additional intron analysis improves the diagnostic yield.[11] A powerful intron analysis tool, spliceAI [13] uses deep residual neural networks to identify splice-relevant mutations or splice mutations that result in aberrant isoforms, is available. SpliceAI could help diagnose rare diseases [14 15] Conversely, whole genome sequencing could be more potent than WES as a powerful genomic diagnostic method for diseases from unknown causes.[16] However, considering various cost aspects, re-analysing WES is still appropriate given its practicality. [17]

Therefore, we hypothesised that the re-evaluation WES data using the LOEUF score and additional intron analysis would improve the diagnostic yield of WES and identify new causal genes in cases of foetal anomalies. We aimed to investigate the cause of foetal morbidity in 138 cases from 2011 to 2021 using WES and copy number variation (CNV) analyses and find new disease-causing genes among them. For undiagnosed cases after conventional WES method, we employed three additional approaches including intron analysis in combination with the LOEUF score using WES data.

## METHODS

### Study design and participants

This study involved 138 cases of foetal structural abnormalities and we performed trio comprehensive genomic analyses on patients and their parents at the National Centre for Child Health and Development between 2011 and 2021. The participants exhibited a structural anomaly during a prenatal ultrasound (after 11 weeks of gestation) that was detected at multiple perinatal centres.

After evaluations by foetal imaging specialists and clinical genetic experts, samples from participants requiring WES for diagnosis were sent to one tertiary-level referral centre (National Centre for Child Health and Development, Tokyo, JAPAN). Foetuses with suspected apparent chromosomal abnormalities or with a known infection were excluded. Although some cases had increased nuchal translucency, participants with only increased nuchal translucency were not included. The study was approved by the Institutional Review Board (IRB) of the National Centre for Child Health and Development, Japan (IRB number: 236, 926). Written informed consent was obtained from the patient or their legal representatives for this research work. Based on the IRB-approved research plan, a trio genomic analysis was conducted with the informed consent of the patients and their families; specimens were further obtained in the perinatal period (foetal and neonatal).

### Comprehensive genetic analysis (WES and CNV analyses)

Genomic DNA was collected from the cord and peripheral blood, umbilical and placenta, amniotic fluid cells, and saliva. The analysis—which was first performed in 2011—was refined again in March 2022 using the latest pipeline for all families. Following the manufacturer’s protocol, a whole-exome library of each sample was prepared using either the SureSelect v5 (early cases) or v6 (later cases) Capture Kit (Agilent Technologies, CA, USA). The libraries were sequenced on HiSeq 2500 (Illumina, CA, USA) using the 101-or 150-bp paired-end mode for samples obtained before 2019. The software programs used were HiSeq Control Software v2.2.68, Real-Time Analysis (RTA) v1.18.66.3, and bcl2fastq v1.8.4 to prepare fastq files. For samples obtained after 2019, the library was prepared using the SureSelect v6 Capture Kit (Agilent Technologies) and sequenced using a DNBSEQ-G400 BGI sequencer (BGI, Shenzhen, China). Mutational signature analysis was performed using the Burrows-Wheeler Aligner (BWA)-MEM[18]/Genome Analysis Toolkit (GATK) HaplotypeCaller[19] for variant analysis. Thereafter, sequence reads were mapped and aligned to the reference genome sequence hs37d5 using our pipeline, which included GrepWalk(https://aihospital.ncchd.go.jp/grepwalk/), Cutadapt v2.6[20], BWA v0.7.15[21], SAMtools v1.6[22], Picard v2.1.1[23], GATK v3.7[19], and JDK v1.8.0_112. Multi-sample calling of single nucleotide variants (SNVs) and short indels was performed using the RefSeq gene database in combination with eight in-house control datasets from adult women—who had not experienced abnormal pregnancies or deliveries—as the control. The variant frequencies were compared to reference sets from the Integrated Japanese Genome Variation Database (3.5KJPN). In all cases, we performed CNV analysis using EXCAVATOR 2 v1.1.2, a WES-based CNV detection tool.[24] The CNV analysis of WES data was followed by genome-wide single nucleotide polymorphism (SNP) array analysis that was performed using the Infinium Asian Screening Array-24 v1.0 BeadChip Kit (Illumina). For the causal variants of the narrowed results, we used the American College of Medical Genetics (ACMG) criteria and selected candidate causal variants that were likely pathogenic or pathogenic.[25]

### RNA sequencing

The total RNA was extracted from the placental villi of case #23 and normal control using the RNeasy Fibrous Tissue Mini Kit (Qiagen, Hilden, Germany).Genomic DNA was removed using RNase-Free DNase Set (Qiagen); then, we prepared RNA sequence library using the NEBNext Ultra II Directional RNA Library Prep Kit for Illumina (New England Biolabs, MA, USA), and sequenced on NovaSeq6000 (Illumina) using 100-bp paired-end mode. We carried out mapping using HISAT2.[26]

### Disease type definition and classification

Skeletal dysplasia, cardiovascular, central nervous system, and kidney abnormalities were defined as the only existing structural abnormalities in each organ, respectively. Multiple malformations were defined as the presence of structural abnormalities in more than one organ. Foetal growth abnormality only included foetal growth restriction (FGR). FGR was defined as -1.5 standard deviation or less of the estimated foetal weight without hypertensive disorders of pregnancy. Hydrops fetalis was defined as the presence of abnormal amounts of fluid build-up in two or more body areas of a foetus without any structural anomalies. Effusion was defined as the presence of foetal pleural effusion that required foetal surgical treatment in this study.

### LOEUF score

Genes with fewer functional variants in healthy individuals are more likely to cause disease than genes with more functional variants. If a gene has relatively less functional variations than that expected in the general population, it is considered ‘intolerant’. Based on the WES data of 125,748 individuals from the Genome Aggregation Database, version 2.1.1, the degree of intolerance of each gene and predicted LOF variation were directly assessed using a continuous measure of the observed/predicted ratio. Subsequently, the confidence interval of the ratio was estimated, and the 90% upper limit of this confidence interval was scored as LOEUF.[8]

### Re-evaluation on known disease-causing genes

For cases that remained undiagnosed with our conventional WES method, in the second step, we made known disease-causing gene panels and re-evaluated variants including introns on these genes using spliceAI. We prepared gene lists and re-analysed WES data on these genes for hydrops fetalis, skeletal disorder, Joubert syndrome, and Meckel–Gruber syndrome (Supplemental Table 2), because these diseases were included in the clinical diagnosis in many cases of our study. In this approach we used spliceAI with default setting on a web browser when suspicious pathogenic variants were identified.

### Comprehensive intron analysis pipeline using spliceAI among 2,971 genes with a low LOEUF score

We prepared files in General Transfer Format (GTF) after variant calling using haplotypecaller and a bed file containing information on the chromosome number, start, and end of the 2,971 genes with a LOEUF score, which was 0.35 or less, and selected only the variants on that gene. Next, we selected the variants without a reference SNP (rs) number and ran spliceAI for these variants on the command line with the default settings and hs37d5 as the reference genome sequence data. From the obtained GTF files, the pathogenic variants were narrowed down as targets of autosomal dominant diseases with *de novo*, X-linked dominant (XLD), or X-linked recessive disorders (XLR), as a loss of function in genes with a low LOEUF score could lead to haploinsufficiency.

### LOEUF score comparison of OMIM database genes

All 17,425 listed genes in the OMIM (Online Mendelian Inheritance in Man, https://www.omim.org/downloads), were divided into autosomes and X chromosomes, and genes with LOEUF scores were further selected. The number of autosomal genes and X-chromosomal genes having LOEUF score was 13,716 and 593, respectively. The genes were further classified according to the inheritance form of the disease based on the phenotype information registered in the OMIM. The LOEUF scores were compared for each category.

### Statistical analyses

Welch’s *t*-test was performed to compare the LOEUF scores. Statistical analysis was performed using R v4.1.0 (R Foundation for Statistical Computing, Vienna, Austria).

## RESULTS

Figure 1 shows systematic diagnostic method and diagnosed cases with WES in this study.

**Figure 1.**
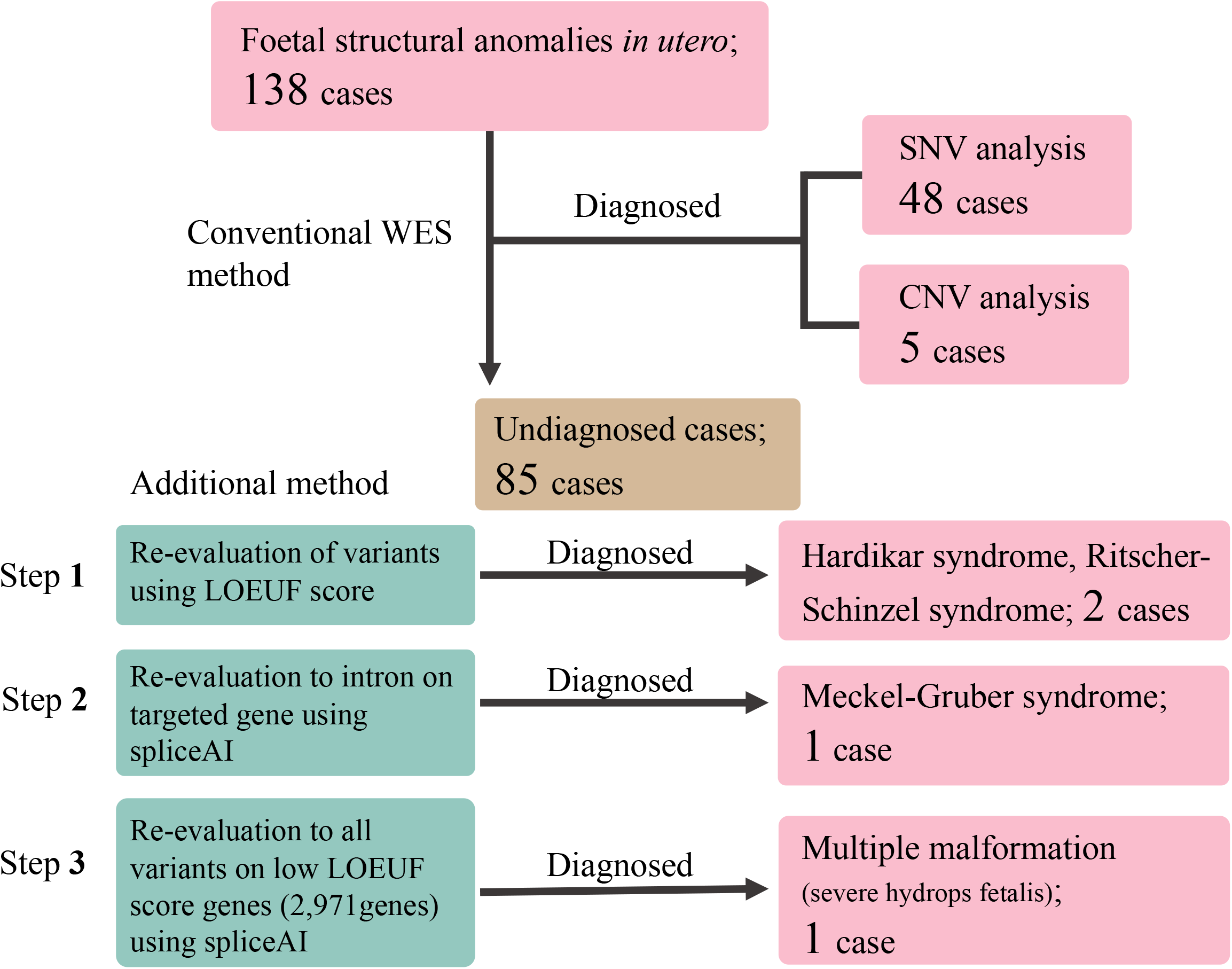
Diagnostic method and diagnosed cases on WES.

**Figure 2.**
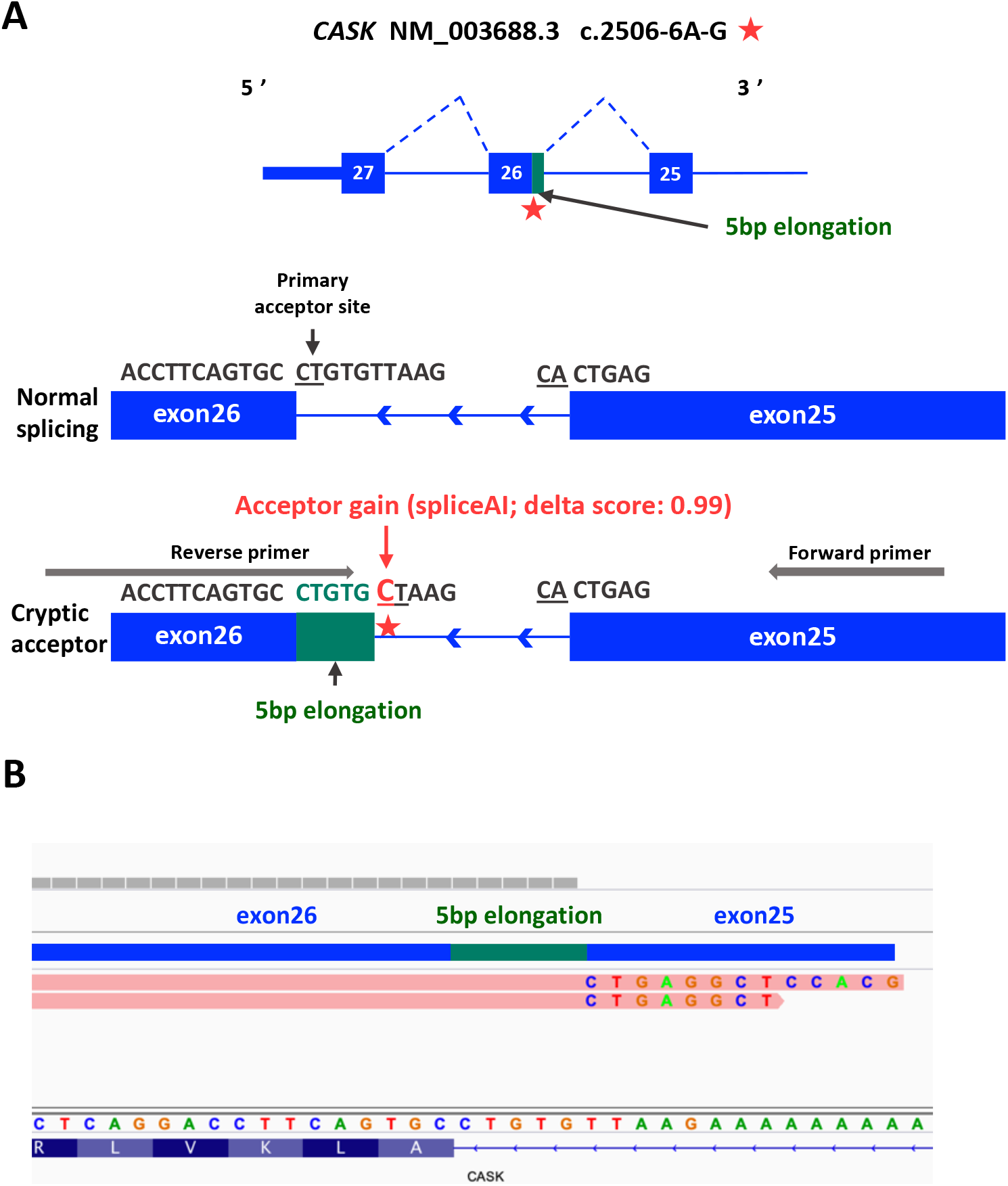
Aberrant splicing of *CASK*. **(A)** The overview of the intronic variant lead aberrant splicing. (**B**) RNA sequencing showed aberrant splicing on CASK.

### Diagnostic results with the conventional method

Causative pathological variants of SNVs or CNV were identified in 53 of the 138 cases (38.4%) using the conventional WES method. As we were accordingly re-evaluating each case and adapting the new database as appropriate, none of the new diagnoses were in the latest conventional WES pipeline. Details of all cases are presented in Supplemental Table 1. Table 1 shows the number of cases and diagnostic yields by outcome and phenotypic classification using the conventional WES method. The diagnostic yield for cardiovascular abnormalities and skeletal dysplasia was high (Table 1).

**Table 1.**
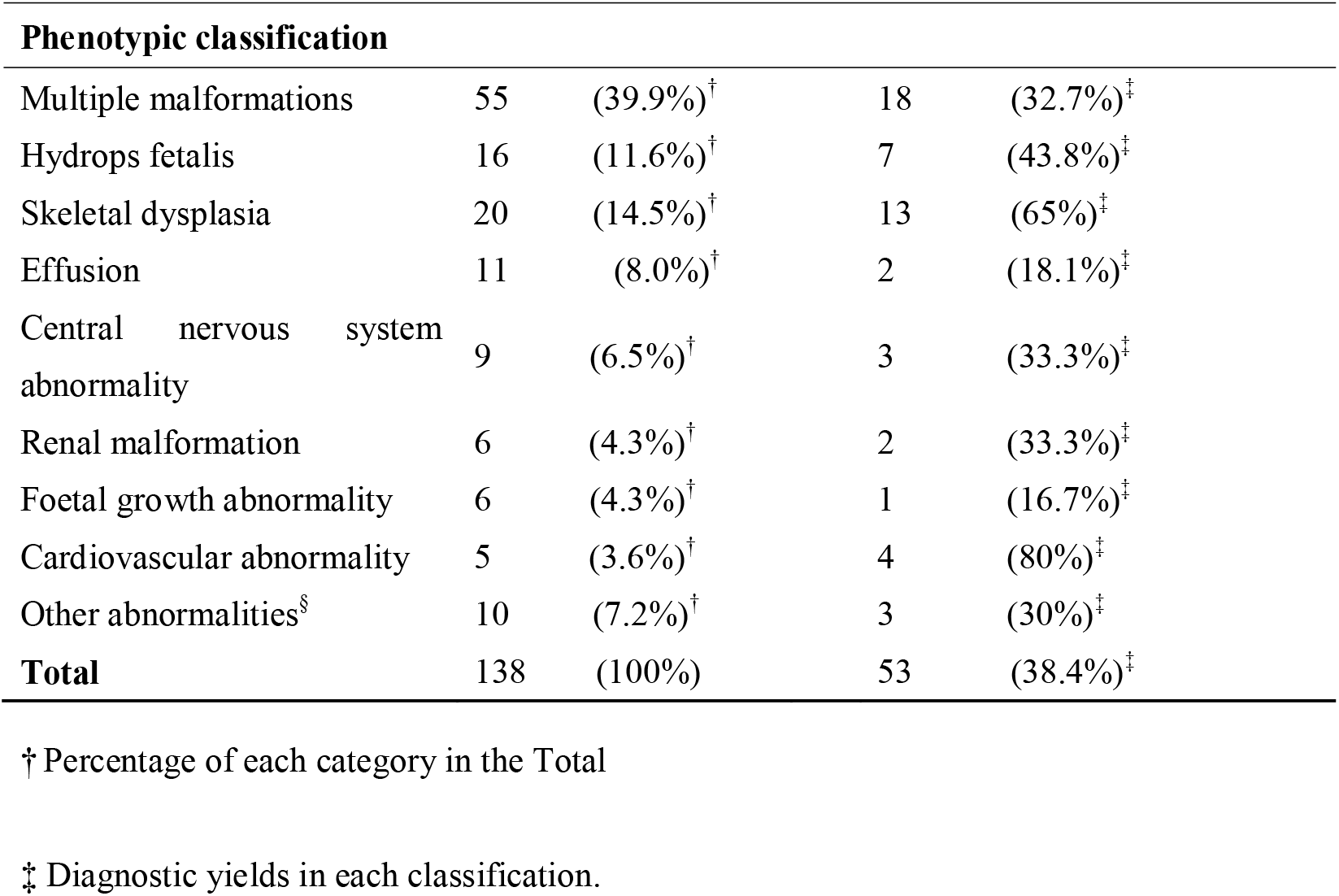
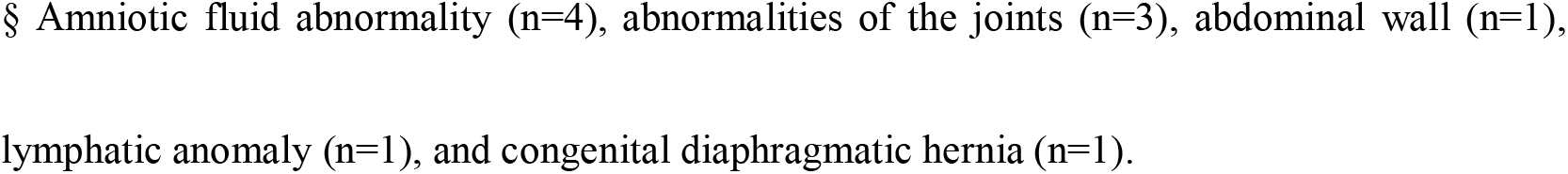
Breakdown of phenotypic classification in 138 cases with conventional WES analysis.

| **Phenotypic classification** |  |  |  |  |
| --- | --- | --- | --- | --- |
| Multiple malformations | 55 | (39.9%)<sup>†</sup> | 18 | (32.7%)<sup>‡</sup> |
| Hydrops fetalis | 16 | (11.6%)<sup>†</sup> | 7 | (43.8%)<sup>‡</sup> |
| Skeletal dysplasia | 20 | (14.5%)<sup>†</sup> | 13 | (65%)<sup>‡</sup> |
| Effusion | 11 | (8.0%)<sup>†</sup> | 2 | (18.1%)<sup>‡</sup> |
| Central nervous system abnormality | 9 | (6.5%)<sup>†</sup> | 3 | (33.3%)<sup>‡</sup> |
| Renal malformation | 6 | (4.3%)<sup>†</sup> | 2 | (33.3%)<sup>‡</sup> |
| Foetal growth abnormality | 6 | (4.3%)<sup>†</sup> | 1 | (16.7%)<sup>‡</sup> |
| Cardiovascular abnormality | 5 | (3.6%)<sup>†</sup> | 4 | (80%)<sup>‡</sup> |
| Other abnormalities<sup>§</sup> | 10 | (7.2%)<sup>†</sup> | 3 | (30%)<sup>‡</sup> |
| **Total** | **138** | **(100%)** | **53** | **(38.4%)<sup>‡</sup>** |

<sup>†</sup> Percentage of each category in the Total
<sup>‡</sup> Diagnostic yields in each classification.
§ Amniotic fluid abnormality (n=4), abnormalities of the joints (n=3), abdominal wall (n=1), lymphatic anomaly (n=1), and congenital diaphragmatic hernia (n=1).

### CNV analysis results

Along with SNV analysis, we performed CNV analysis using WES data or the SNP array. We identified the cause of the disease in five cases. Trisomy 21, Monosomy X, and Trisomy 18 were identified in one case each (cases #4, #22, and #34, respectively). The phenotypes of these popular chromosomal abnormality cases were so complex that even foetal imaging specialists could not suspect them clinically. Case #26 was diagnosed with FGR and thoracic hypoplasia; however, the molecular genetic diagnosis was Sotos syndrome by del(5)(q35.2→q35.3) of 1.9 Mb containing *NSD1*. This might have been caused by a discrepancy in the gestational age due to a mistake in determining the expected delivery date. In case #101, we identified 12-Mb deletions (del(7)(q35→qter)) after CNV evaluation using WES data. These deletions are reported pathogenic deletions.[27]

### Additional intron analysis in combination with genetic intolerance score for undiagnosed cases

For 85 cases that remained undiagnosed with our conventional WES pipeline, we performed a re-analysis of WES data using three additional steps to diagnose. We re-evaluated 1) candidate variants using a LOEUF score, 2) all variants on disease-causing genes using spliceAI, and 3) the rare variants in all low LOEUF scored genes (< 0.35) using spliceAI. (Figure 1).

### First step: Re-evaluation of candidate variants using the LOEUF score

Case #24 had multiple malformation, including left cleft lip and palate, large ventricular septal defect, diaphragmatic hernia, left hydronephrosis and hydroureter, and omphalocele. The phenotype was complicated to diagnose genetically using the conventional WES method, although one *de novo* variant on the splice acceptor site of *MED12* is in the final candidate variant list. However, the LOEUF score could help us re-evaluate the variant on *MED12*, whose LOEUF score is extremely low (0.071) possibly causing haploinsufficiency. Based on a recent case report with similar specific phenotype, we diagnosed the patient to have Hardikar syndrome, an XLD disease, with a diaphragmatic hernia.[28]

Case #54 had multiple malformation syndrome, including scalp defects, nasal root hypoplasia, micrognathia, thoracic hypoplasia, unilateral cryptorchidism, slightly short limbs and ribs, and toe syndactyly; however, we could not identify any genes caused by specific monogenic diseases. Using the LOEUF score, a pathogenic *CCDC22* variant could be identified (LOEUF score: 0.123). *CCDC22*-associated Ritscher-Schinzel syndrome type 2 is inherited in an XLR pattern.[29] However, as there were no major characteristic symptoms of Ritscher-Schinzel syndrome, such as cardiac and intracranial anomalies, the clinical diagnosis was difficult because this case had only minor symptoms.

### Second step: Re-evaluation of variants including intron on known disease-causing genes

We tried to re-evaluate WES data of undiagnosed hydrops fetalis and effusion cases (18 cases), skeletal disorders (seven cases), Joubert syndrome (one cases), and Meckel–Gruber syndrome (two cases) using spliceAI on these disease-causing genes. Although case #129 was clinically diagnosed to have Meckel–Gruber syndrome, an autosomal recessive disease, we could only detect one stop gain *TMEM67* variant with the conventional WES method. Evaluating all variants including introns on *TMEM67*, we identified a likely pathogenic variant (c.2100+3A>G). spliceAI predicted splice donor loss (delta score by spliceAI; 0.38). MaxEntScan[30] also predicted a broken conventional splice donor site (5.96>-1.06 (−117.79%)). This variant might cause aberrant splicing, e.g. exon deletion, which leads to the translation of truncated proteins.

### Third step: Comprehensive intron analysis in combination with a low LOEUF score

To find overlooked causative variants, we re-evaluated the candidate variants, including the intronic region in all low LOEUF score genes (less than 0.35) using spliceAI (see Methods). We could identify one *de novo* hemizygous pathogenic variant that could cause aberrant splicing on *CASK* in case #23. Foetal ultrasonography showed severe hydrops fetalis, polyhydramnios, and mild cerebellar hypoplasia. *CASK* mutation caused mental retardation and microcephaly with pontine and cerebellar hypoplasia (MICPCH) syndrome, an XLD disease. The LOEUF score of *CASK* is 0.073, and the acceptor-gain delta score of the intron variant is 0.99 using spliceAI. RNA sequencing revealed that this intronic variant lead to aberrant splicing and a 5-bp gain elongation of the intronic sequence, which could cause haploinsufficiency of *CASK*(FigureA,B).

### X-linked disease-causing genes are intensely intolerant

In our study, all X-linked disorder genes had LOEUF scores < 0.2 (Figure 3A), suggesting that the LOEUF score could help narrow down the candidates for X-linked disorder genes, especially foetal-onset diseases. Therefore, we determined the LOEUF scores of all disease-causing genes from the OMIM and listed the causative genes (see Methods); the LOEUF scores could be determined for 14,309 causative genes (Supplemental Table 3). The LOEUF scores of each X-linked gene were significantly lower than those of autosomal-dominant genes (Figure 3B), suggesting that X-linked disease-causing genes are strongly intolerant. Therefore, the LOEUF scores of the disease-causing genes in X-linked disorders might help narrow down the causative genes.

**Figure 3.**
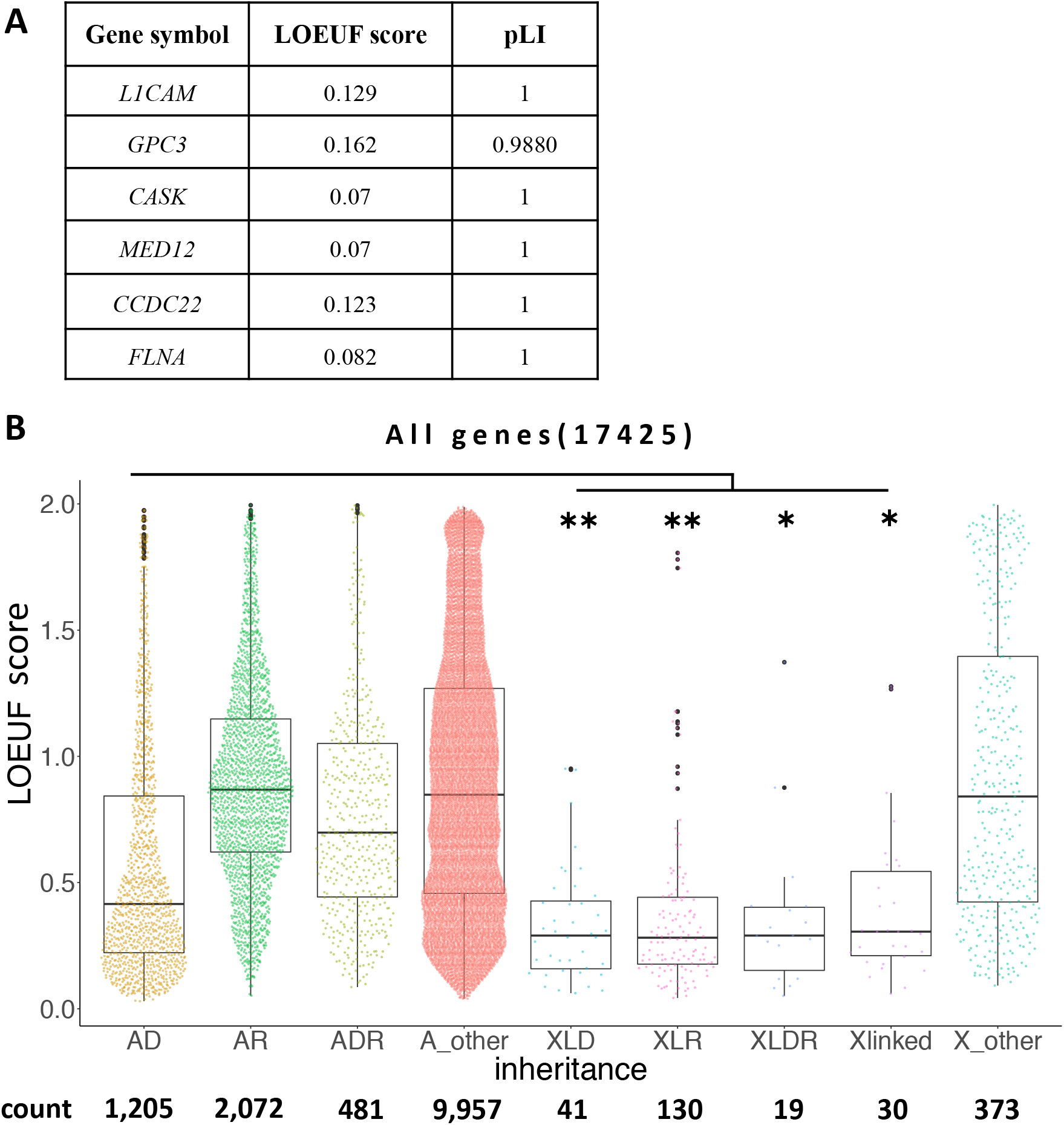
Comparison among all OMIM registered genes. **(A)** The list of LOEUF score of genes on chromosome X, diagnosed in our study. (**B**) The comparison among all OMIM registered genes. \**P* < 0.001. \*\**P* < 0.001 LOEUF loss-of-function observed/expected upper bound fraction; AD, autosomal dominant; AR, autosomal recessive; ADR, autosomal dominant and recessive; A_other, other genes of autosome; XLD, X-linked dominant; XLR, X-linked recessive; XLDR, X-linked dominant and recessive; X_other, other genes of chromosome X.

## DISCUSSION

In 138 cases of unexplained foetal structural anomalies, we genetically diagnosed 47 cases (33%) by SNV analysis and five cases by CNV analyses using the conventional WES method. For cases that remained undiagnosed with our conventional WES method, re-analysis including the intronic region in combination with a genetic intolerance score allowed us to evaluate the variants of unknown significance as pathogenic variants, especially on chromosome X. This is the first study to demonstrate that comprehensive intron analysis of WES data in combination with LOEUF scores helps us find causative genes overlooked by conventional WES and one new phenotype of known disease. Through our diagnosed X-linked disorder cases, we could confirm that evaluation using the LOEUF score was considerably more helpful in diagnosing X-linked disorders. Here, we showed that additional intron analysis of WES in combination with LOEUF scores improved diagnostic yields.

Our method also showed the usefulness of WES re-evaluation for the diagnosis of foetal-onset disorders. Some previous studies showed that WES re-analysis is useful for monogenic diseases.[10 11] Elias et al. showed that the latest annotation of WES among likely monogenic rare diseases could improve molecular diagnostic yields by 5.9% compared with the initial analysis.[10] Yingdi et al. tried to re-analyse WES data among 174 undiagnosed possible Mendelian diseases along with the evaluation of CNV, intron regions, and InDels within the 10-50-bp detection method. They might have finally reduced missed diagnoses by 8%.[11] Here, we showed that the re-analysis of WES data adding intron analysis together with genetic intolerance score can improve the diagnostic yield by 2.9% (4/138) including cases with complex phenotypes of foetal morphology in which monogenic disorder is not expected.

We employed the three steps re-evaluation to diagnose undiagnosed cases with our conventional WES method. Finally, we could have four additional diagnoses. In the first step, which focused on the LOEUF score, we could diagnose two X-linked disorders, with complicated phenotypes, by re-evaluating the narrowed-down variants based on the latest literature. In the second step, we showed that intron evaluation of targeted genes was important for undiagnosed monogenic disorders (case #129). Similarly, in one repeated hydrops fetalis case (#123), we detected one stop gain mutation in *PIEZO1* and the whole deletion of *PIEZO1* on another allele. If we had detected one pathogenic variant on targeted genes, it was important to carefully check intron or microdeletion on disease-causing genes. In the third step, we found a new phenotype in X-linked monogenic disorder. We showed that it was possible to obtain marked potential for the diagnosis and identification of new phenotypes by verifying genes that can cause haploinsufficiency, including introns, even in phenotypes that have not been reported to date. Few CASK-related diseases have been reported in detail with symptoms from the foetal period, and almost all cases have been reported to be uneventful during pregnancy. Moreover, even in males, severe hypoplasia of the cerebellum and bridges has not been noted in CASK-related diseases during foetal periods and almost all cases have been reported to be uneventful during pregnancy.[31 32] Although the biological sample was very old, RNA sequencing reveal that this intronic variant had lead aberrant splicing. Clinically, the possibility of gonadal mosaicism cannot be ruled out because of the previous unexplained IUFD. Eventually, this study suggested that CASK-related disease cause severe hydrops fetalis and IUFD even if cerebellar hypoplasia in utero was mild.

As the LOEUF scores of the causative genes detected in all X-linked cases were very low (Figure 3A), we investigated whether the LOEUF scores might be useful for evaluating X-linked diseases. Analysis using all genes from the OMIM revealed remarkably low LOEUF scores of the genes causing X-linked disorders extracted from the OMIM (Figure 3B), indicating that the evaluation of causative variants and genes using LOEUF scores might be helpful, especially on chromosome X. Reportedly, the LOF variants of genes with LOEUF scores < 0.239 are enriched in stillbirth cases, even in non-OMIM genes,[5] which may be helpful to select new causative genes among other populations. Indeed, we showed that systematic re-evaluation focusing on low LOEUF scores helped diagnose and found the new phenotype.

Our study has some limitations. More than half of the cases were undiagnosed, and it was difficult to identify the cause of morphological abnormality in these cases using WES. In the future, it is necessary to use WGS to evaluate SVs other than SNVs, such as indel size, promoter and enhancer dysfunctions, splicing abnormality by deep intronic variants, and translocations. There are approximately 1,500 genes with LOEUF scores < 0.239. Among these, many genes are not yet reported to be associated with human diseases; therefore, we expect to accumulate more data in the future. However, this re-analysis method, using all open-source software, is easy to implement. Furthermore, it is also readily adaptable to WGS data. It may be very useful for additional evaluation of WES data of diseases of unknown origin.

This study showed that re-evaluation of WES data, including introns, in combination with the LOEUF score was useful for diagnosing foetal onset diseases with ambiguous phenotypes, especially in X-liked diseases, to improve the diagnostic yield.

## Data Availability

All data produced in the present study are available upon reasonable request to the authors.

## ACKNOWLEDGEMENTS

We are deeply grateful to the patients and their family members for participating in this study. We would also like to express our gratitude to the attending physicians of the patients and our colleagues in the laboratory for their cooperation in the analysis.

## REFERENCES

1. Lord J, McMullan DJ, Eberhardt RY, et al. Prenatal exome sequencing analysis in fetal structural anomalies detected by ultrasonography (PAGE): A cohort study. Lancet 2019;393(10173):747–57. https://doi.org/10.1016/S0140-6736(18)31940-8.

2. Petrovski S, Aggarwal V, Giordano JL, et al. Whole-exome sequencing in the evaluation of fetal structural anomalies: a prospective cohort study. Lancet 2019;393(10173):758–67. https://doi.org/10.1016/S0140-6736(18)32042-7.

3. Hunter LE, Simpson JM. Prenatal screening for structural congenital heart disease. Nat Rev Cardiol 2014;11(6):323–34. https://doi.org/10.1038/nrcardio.2014.34.

4. Sparks TN, Lianoglou BR, Adami RR, et al. Exome sequencing for prenatal diagnosis in nonimmune Hydrops fetalis. N Engl J Med 2020;383(18):1746–56. https://doi.org/10.1056/NEJMoa2023643.

5. Stanley KE, Giordano J, Thorsten V, et al. Causal genetic variants in stillbirth. N Engl J Med 2020;383(12):1107–16. https://doi.org/10.1056/NEJMoa1908753.

6. Saunders CJ, Miller NA, Soden SE, et al. Rapid whole-genome sequencing for genetic disease diagnosis in neonatal intensive care units. Sci Transl Med 2012;4(154):154ra35. https://doi.org/10.1126/scitranslmed.3004041.

7. Tolusso LK, Hazelton P, Wong B, et al. Beyond diagnostic yield: prenatal exome sequencing results in maternal, neonatal, and familial clinical management changes. Genet Med 2021;23(5):909–17. https://doi.org/10.1038/s41436-020-01067-9.

8. Karczewski KJ, Francioli LC, Tiao G, et al. The mutational constraint spectrum quantified from variation in 141,456 humans. Nature 2020;581(7809):434–43. https://doi.org/10.1038/s41586-020-2308-7.

9. Petrovski S, Wang Q, Heinzen EL, et al. Genic intolerance to functional variation and the interpretation of personal genomes. PLoS Genet 2013;9(8):e1003709. https://doi.org/10.1371/journal.pgen.1003709.

10. Salfati EL, Spencer EG, Topol SE, et al. Re-analysis of whole-exome sequencing data uncovers novel diagnostic variants and improves molecular diagnostic yields for sudden death and idiopathic diseases. Genome Med 2019;11(1):83. https://doi.org/10.1186/s13073-019-0702-2.

11. Liu Y, Teng Y, Li Z, et al. Increase in diagnostic yield achieved for 174 whole-exome sequencing cases reanalyzed 1-2 years after initial analysis. Clin Chim Acta 2021;523:163–68. https://doi.org/10.1016/j.cca.2021.09.015.

12. Dai P, Honda A, Ewans L, et al. Recommendations for next generation sequencing data reanalysis of unsolved cases with suspected Mendelian disorders: A systematic review and meta-analysis. Genet Med 2022;24(8):1618–29. https://doi.org/10.1016/j.gim.2022.04.021.

13. Jaganathan K, Kyriazopoulou Panagiotopoulou S, et al. Predicting splicing from primary sequence with deep learning. Cell 2019;176(3):535–48e24. https://doi.org/10.1016/j.cell.2018.12.015.

14. Lord J, Baralle D. Splicing in the diagnosis of rare disease: advances and challenges. Front Genet 2021;12:689892. https://doi.org/10.3389/fgene.2021.689892.

15. Setty ST, Scott-Boyer MP, Cuppens T, et al. New developments and possibilities in reanalysis and reinterpretation of whole exome sequencing datasets for unsolved rare diseases using machine learning approaches. Int J Mol Sci 2022;23(12):241. https://doi.org/10.3390/ijms23126792.

16. Stranneheim H, Lagerstedt-Robinson K, Magnusson M, et al. Integration of whole genome sequencing into a healthcare setting: high diagnostic rates across multiple clinical entities in 3219 rare disease patients. Genome Med 2021;13(1):40. https://doi.org/10.1186/s13073-021-00855-5.

17. Ewans LJ, Minoche AE, Schofield D, et al. Whole exome and genome sequencing in mendelian disorders: a diagnostic and health economic analysis. Eur J Hum Genet 2022;30(10):1121–31. https://doi.org/10.1038/s41431-022-01162-2.

18. Li H. Aligning sequence reads, clone sequences and assembly contigs with BWA-MEM. arXiv [Preprint]. 2013. https://doi.org/10.48550/arXiv.1303.3997.

19. Poplin R, Valentin R-R, DePristo Ma, et al. Scaling accurate genetic variant discovery to tens of thousands of samples. bioRxiv [Preprint]. 2018. https://doi.org/10.1101/201178.

20. Martin. M. Cutadapt removes adapter sequences from high-throughput sequencing reads. EMBnet j.;17(1). https://doi.org/10.14806/ej.17.1.200.

21. Li H, Durbin R. Fast and accurate short read alignment with Burrows-Wheeler transform. Bioinformatics 2009;25(14):1754–60. https://doi.org/10.1093/bioinformatics/btp324.

22. Li H, Handsaker B, Wysoker A, et al. The sequence alignment/map format and SAMtools. Bioinformatics 2009;25(16):2078–9. https://doi.org/10.1093/bioinformatics/btp352.

23. Broad Institute. Picard Toolkit. 2019 (accession date: 4th May 2016). https://broadinstitute.github.io/picard/.

24. D’Aurizio R, Pippucci T, Tattini L, et al. Enhanced copy number variants detection from whole-exome sequencing data using EXCAVATOR2. Nucleic Acids Res 2016;44(20):e154. https://doi.org/10.1093/nar/gkw695.

25. Richards S, Aziz N, Bale S, et al. Standards and guidelines for the interpretation of sequence variants: a joint consensus recommendation of the American College of Medical Genetics and Genomics and the Association for Molecular Pathology. Genet Med 2015;17(5):405–24. https://doi.org/10.1038/gim.2015.30.

26. Pertea M, Kim D, Pertea GM et al. Transcript-level expression analysis of RNA-seq experiments with HISAT, StringTie and Ballgown. Nat Protoc 2016;11(9):1650–67. https://doi.org/10.1038/nprot.2016.095.

27. Schinzel A. Catalogue of unbalanced chromosome aberrations in man. 2nd rev. and expanded ed. Berlin ; New York: Walter de Gruyter, 2001:317–8.

28. Li D, Strong A, Shen KM, et al. De novo loss-of-function variants in X-linked MED12 are associated with Hardikar syndrome in females. Genet Med 2021;23(4):637–44. https://doi.org/10.1038/s41436-020-01031-7.

29. Kolanczyk M, Krawitz P, Hecht J, et al. Missense variant in CCDC22 causes X-linked recessive intellectual disability with features of Ritscher-Schinzel/3C syndrome. Eur J Hum Genet 2015;23(5):633–8. https://doi.org/10.1038/ejhg.2014.109.

30. Yeo G, Burge CB. Maximum entropy modeling of short sequence motifs with applications to RNA splicing signals. J Comput Biol 2004;11(2-3):377–94. https://doi.org/10.1089/1066527041410418.

31. Najm J, Horn D, Wimplinger I, et al. Mutations of CASK cause an X-linked brain malformation phenotype with microcephaly and hypoplasia of the brainstem and cerebellum. Nat Genet 2008;40(9):1065–7. https://doi.org/10.1038/ng.194.

32. Gafner M, Boltshauser E, D’Abrusco F, et al. Expanding the natural history of CASK-related disorders to the prenatal period. Dev Med Child Neurol 2022. https://doi.org/10.1111/dmcn.15419.

